# Antimicrobial Resistance Gene Distribution and Population Structure of *Escherichia coli* isolated from Humans, Livestock, and the Environment: Insights from a One Health Approach

**DOI:** 10.1101/2025.04.07.25325408

**Authors:** Beatus Lyimo, Valery Sonola

**Affiliations:** School of Life Sciences and Bioengineering, Nelson Mandela African Institution of Science and Technology, Arusha; Livestock Training Agency (LITA), Buhuri Campus, P. O. Box 1483, Tanga, Tanzania

**Keywords:** *Escherichia coli*, antimicrobial resistance, One Health, genomic surveillance, plasmids, zoonotic transmission

## Abstract

Antimicrobial resistance (AMR) is an escalating public health threat, with evidence highlighting the exchange of resistance genes among humans, animals, and the environment. Whole-genome sequence (WGS) offers high-resolution pathogen subtyping and provides extensive insights into AMR’s early emergence and spread. This study investigates the distribution of AMR genes, plasmid types, and the population structure of Escherichia coli isolates from humans, livestock, fish, and the environment. A total of 244 WGS datasets of E. coli isolates were pulled from a public database from multiple studies and analyzed to characterize AMR gene distribution, plasmid diversity, and population structure across humans, livestock and the environment.

The findings reveal widespread dissemination of AMR genes across all sources. Aminoglycoside resistance genes (aac(3)-IId, aph(3’’)-Ib, aph(6)-Id, aadA1, aadA5) and β-lactam resistance genes (bla_TEM-1_, bla_OXA-1_, bla_CTX-M-15_) were prevalent across all environments. Quinolone resistance mutations (gyrA_S83L, gyrA_D87N, parC_S80I) were also shared among human, livestock, fish, and environmental isolates, indicating cross-species transmission. Tetracycline resistance genes (tet(A), tet(B), tet(D)) were found in humans, livestock, and fish. Plasmid types IncFIA, IncI1, and IncFII exhibited extensive cross-source sharing, with strong connectivity between humans and livestock. Principal Component Analysis (PCA) revealed that E. coli isolates from Kenya formed a tight, distinct cluster, while others were more dispersed. The Minimum Spanning Tree (MST) network showed the clusters where human and livestock isolates were closely connected, it further showed some human isolates cluster with fish and environmental isolates. The MST network demonstrated close clustering of human and livestock isolates, indicating possible cross-species transmission. These findings showed the interconnected nature of AMR across human, animal, and environmental sectors and underscored the need for integrated surveillance under a One Health framework to monitor and control the spread of clinically significant AMR genes.

## Background

Antimicrobial Resistance (AMR) is one of the biggest global public health threats as it is the leading cause of death globally, however, its magnitude is not well known (Murray et al., 2022a). In the year 2019, it’s estimated that over 4.95 million deaths have been associated with AMR, including 1.27 million deaths attributed to bacterial infections (Murray et al., 2022a). The impact of AMR-related morbidity and mortality is particularly severe in low- and middle-income countries (LMICs), where individuals are 1.5 times more likely to die from AR infectious organisms compared to those in high-income countries. This disparity is even more pronounced among young children. Among children under the age of five who die from AR infections, 99.65% are in low- or middle-income countries (Baker et al., 2018; UNICEF, 2023). In 2019, the United Republic of Tanzania reported an estimated 12,500 deaths directly attributable to AMR and an additional 54,000 deaths associated with AMR-related infections (Murray et al., 2022a). Moreover, Tanzania was ranked as the 30th highest globally in terms of age-standardized mortality rates associated with AMR, among 204 countries analyzed in a global study (Murray et al., 2022). In 2016, Tanzania formulated a National Action Plan for AMR (2017–2022) in response to recommendations from the WHO and the Global Health Security Agenda Joint External Evaluation (Neema et al., 2023; WHO, 2015). Following this, a comprehensive One Health AMR Surveillance Framework was developed to facilitate the establishment of AMR surveillance systems across human, animal, and environmental health sectors (United Republic of Tanzania, 2018).

The 2022 Global Burden of Disease study highlighted *Escherichia coli*, *Staphylococcus aureus*, *Klebsiella pneumoniae*, *Streptococcus pneumoniae*, *Acinetobacter baumannii*, and *Pseudomonas aeruginosa* as the six primary contributors to AMR-related deaths, accounting for 73% of such fatalities in 2019 (Murray et al., 2022b). Among these, *E. coli* is often used as a standard indicator of water contamination, reflecting faecal pollution and the potential presence of other harmful microorganisms. While many strains of *E. coli* are harmless and part of the normal flora, however under favorable conditions, they can cause infections such as gastroenteritis, urinary tract infections (UTIs), and sepsis when conditions allow. The burden of infections caused by multidrug-resistant *E. coli* is further exacerbated by the combination of factors, including the widespread misuse of antibiotics, poor sanitation infrastructure, and limited access to adequate healthcare services (Lyimo, et al., 2016a; Subbiah et al., 2020). *E. coli* can persist in the gut or other body sites without causing disease and often serve as a reservoir for potential infection. Under certain conditions, such as immune suppression or body microbiome disruptions due to antibiotics overuse, the colonized *E. coli* strains can change from a harmless state to pathogenic ones. Additionally, the presence of multidrug resistant *E. coli* in the environment particularly in water sources, food, and hospital settings contribute to the infections risk.

Global collective action is required to strengthen AMR surveillance, promote the development of rapid diagnostic tools, and integrate advanced technologies such as WGS to minimize the unnecessary use of antibiotics. The environmental microbiology approach is critical in this effort, as AMR spreads across human, animal, and environmental interfaces, necessitating coordinated interventions. WGS has transformed the study of bacterial pathogens by providing comprehensive insights into phylogenetic relationships, the presence of antimicrobial resistance genes (ARGs), and the detection of virulence (Hazen et al., 2018; Manyahi et al., 2014). The most common extended-spectrum beta-lactamase (ESBL) genes, including bla*_CTX-M_*, bla*_TEM_*, and bla*_SHV_*, confer resistance to third-generation cephalosporins and are frequently associated with mobile genetic elements such as plasmids, enabling their rapid dissemination across different hosts and environments (Nordmann et al., 2011). Integrating One Health principles into AMR surveillance enhances the ability to track resistance patterns, identify transmission pathways, and develop targeted strategies to mitigate its impact on public health, veterinary medicine, and environmental ecosystems (Accogli et al., 2013; Bailey et al., 2011; Peirano et al., 2010; Wu et al., 2014). Although carbapenems are considered last-resort antibiotics, molecular surveillance within a One Health framework has revealed the widespread presence of carbapenemase-producing strains across human, animal, and environmental reservoirs. Studies have identified strains carrying carbapenemase genes, such as *bla_NDM_* and *bla_KPC_* (Hazen et al., 2018; Manyahi et al., 2022; Nordmann & Poirel, 2005) highlighting the urgent need for integrated AMR monitoring and control strategies. The detection of these resistance genes (**Table 1**) emphasizes the necessity of a coordinated approach across public health, veterinary medicine, and environmental sectors to track transmission dynamics and develop effective mitigation strategies (Cella et al., 2023; McEwen & Collignon, 2018; Velazquez-Meza et al., 2022; Welch et al., 2007). One of the most significant contributions of WGS and bioinformatics tools is the ability to trace the movement of antibiotic resistance genes between different bacterial populations. The WGS analysis has shown that resistance genes are often carried on plasmids, integrons, and transposons, which can be transferred between commensal and pathogenic strains. This transfer increases the genetic diversity of resistant *E. coli* strains, complicating efforts to control their spread.

**Table 1:** Summary of major classes of antibiotics, AMR-related genes, and proteins associated with *E. coli*.

| Antibiotic Class | AMR-Related Genes | Associated Proteins | References |
| --- | --- | --- | --- |
| Beta-lactams | *bla<sub>TEM</sub>*, *bla<sub>CTX-M</sub>*, *bla<sub>SHV</sub>*, *bla<sub>OXA</sub>* | Beta-lactamases (e.g., TEM, CTX-M, SHV, OXA) | (Bush et al., 2011; Doi & Paterson, 2007) |
| Aminoglycosides | *aadA*, *aph*, *strA*, *strB* | Aminoglycoside-modifying enzymes (e.g., AadA) | (Ramirez & Tolmasky, 2010) |
| Fluoroquinolones | *gyrA*, *gyrB*, *parC*, *pare* | DNA gyrase and topoisomerase IV | (Ruiz, 2003) |
| Tetracyclines | *tetA*, *tetB*, *tetC* | Efflux pumps (e.g., TetA/B proteins) | (Roberts, 2005) |
| Sulfonamides | *sul1*, *sul2*, *sul3* | Dihydropteroate synthase (Sul proteins) | (Sköld, 2000) |
| Chloramphenicol | *cat*, *cmlA* | Chloramphenicol acetyltransferase (Cat protein) | (Schwarz et al., 2004) |
| Macrolides | *mphA*, *ermB* | Macrolide-modifying enzymes (e.g., MphA) | (Leclercq & Courvalin, 2002) |
| Carbapenems | *bla<sub>KPC</sub>*, *bla<sub>NDM</sub>*, *bla<sub>VIM</sub>*, *bla<sub>OXA-48</sub>* | Carbapenemases (e.g., KPC, NDM, VIM, OXA-48) | (Nordmann et al., 2011) |

This diversity is driven by the exchange of genetic material between human, animal, and environmental strains. In rural areas, zoonotic transmission plays a key role in shaping the genetic landscape of *E. coli*, as livestock are frequently in close contact with humans, allowing for the exchange of bacterial strains and resistance genes. This paper explores the use of the One Health approach to WGS data available in public repositories, focusing on *E. coli* strains circulating in East Africa to examine the distribution of antibiotic resistance genes and genetic diversity of *E. coli* strains across human, animal, and environmental reservoirs.

## Methods

A literature search was conducted across various scientific journals and databases using key terms such as “antibiotic resistance in Tanzania,” “antibiotic resistance in humans in Tanzania,” “antibiotic resistance in livestock in Tanzania,” and “antibiotic resistance in the environment (soil and water).” In each relevant paper, accession number(s) were identified and used to download whole genome sequences (WGS) from public repositories for analysis. All raw WGS data, along with the reference *E. coli* strain K-12 substr. MG1655 were retrieved from the NCBI GenBank database using the accession numbers listed in **Table 2**.

**Table 2.** presents the list of accession numbers for the genomic sequences included in this study. These accession numbers correspond to the *E. coli* isolates analyzed across various sources, including human, livestock, and environmental samples.

| SN | Accession | Reference |
| --- | --- | --- |
| 1 | PRJEB12361 | (Moremi et al., 2016) |
| 2 | PRJEB12376 | (Mshana et al., 2016) |
| 3 | PRJEB71714 | (Kanje et al., 2024) |
| 4 | PRJEB32607 | (Muloi et al., 2022) |

### Data Preprocessing

All data were inspected for quality using fastaqc v0.12.1 to generate individual reports and MultiQC to put all reports together, then trimmed to remove low-quality sequences and adapters using Trimmomati(Bolger et al., 2014)c before downstream analysis.

#### Genome Assembly

Paired-end reads were assembled into contigs using SPAdes. Genome annotation was performed by the Prokka software (Seemann, 2014). Single nucleotide polymorphism (SNP) variant calling was performed using bcftools software (Danecek et al., 2021). In brief, the sequences were mapped to the reference genome *E. coli* str. K-12 substr.MG1655 to generate SAM files and then converted to BAM files and then sorted to produce sorted BAM file, then generate the mpileup. The bcftools was then used for variant calling to generate VCF files of each sample. The individual VCF files were merged to produce one merged vcf file using bcftools.

### AMR Gene Identification

The spade software was used to generate a contig sequence from each pared end sequence. The ResFinder plus software (Florensa et al., 2022) was used to identify AMR genes. In-house Python script followed by pheatmap function in R software was used to generate heatmap plots. The ggVennDiagram function in R was used to show the distribution of shared and unique antimicrobial resistance genes, ARGs) among five different sources.

### Population structure

To assess gene flow between *E. coli* isolated from humans, livestock and environment (soil and water), genetic differentiation was first estimated using the Wright Fixation index (FST) using Vcftools v0.1.5(Danecek et al., 2011) and population structure was determined using principal component analysis (PCA) as implemented in PLINK1.9(Purcell et al., 2007).

### Analysis of Genetic Similarity Among Antimicrobial-Resistant Strains Isolated from Humans, Livestock, Fish, and the Environment

The genetic similarity of antimicrobial-resistant isolates was analyzed using Multilocus Sequence Typing (MLST) v2.23.0, which was downloaded and installed in a Linux environment. The results were then plotted in R using the ggraph and tidygraph packages to visualize genetic relationships among the isolates.

## Results

### AMR Gene Identification and Distributions

A total of 244 WGS sequences were obtained from the NCBI database and analyzed, results showed the distribution of AMR genes across four key sources with several resistance genes found in multiple sources, indicating possible horizontal gene transfer and the movement of resistant bacteria between ecosystems (**Table 3**). Aminoglycoside resistance genes (*aac(3)-IId*, *aph(3’’)-Ib*, *aph(6)-Id*, *aadA1*, *aadA5*) were frequently detected across environmental, fish, human and livestock samples. Similarly, β-lactam resistance genes (bla*_TEM-1_*, bla*_OXA-_1*, bla*_CTX-M-15_*) were identified in all sources, reflecting the widespread dissemination of β-lactamase-producing bacteria. Notably, *bla_CTX-M-15_*, an extended-spectrum β-lactamase (ESBL) gene, was present in humans, livestock, fish and the environment, raising concerns about its impact on clinical treatment options. Quinolone resistance genes (*gyrA_S83L*, *gyrA_D87N*, *parC_S80I*) were also shared across all sources, suggesting cross-species transmission and potential selection pressure from extensive fluoroquinolone use. Sulfonamide resistance genes (*sul1*, *sul2*) were consistently found in all environments, indicating sustained exposure, likely driven by veterinary and agricultural practices. Tetracycline resistance genes (*tet(A)*, *tet(B)*, *tet(D)*) were detected in humans, livestock, and fish, emphasising the global influence of tetracycline use in animal husbandry and its role in environmental contamination.

**Table 3:**
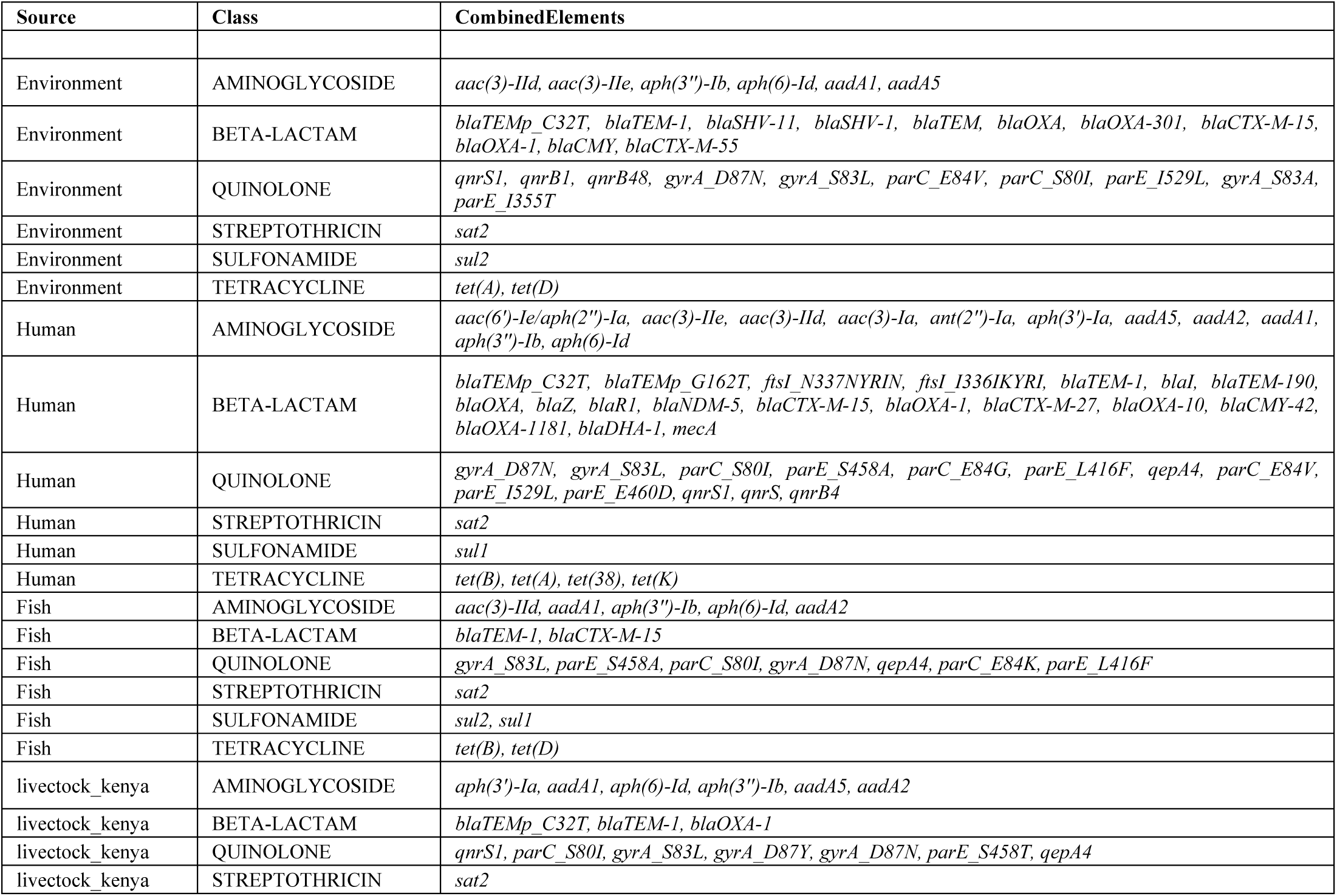
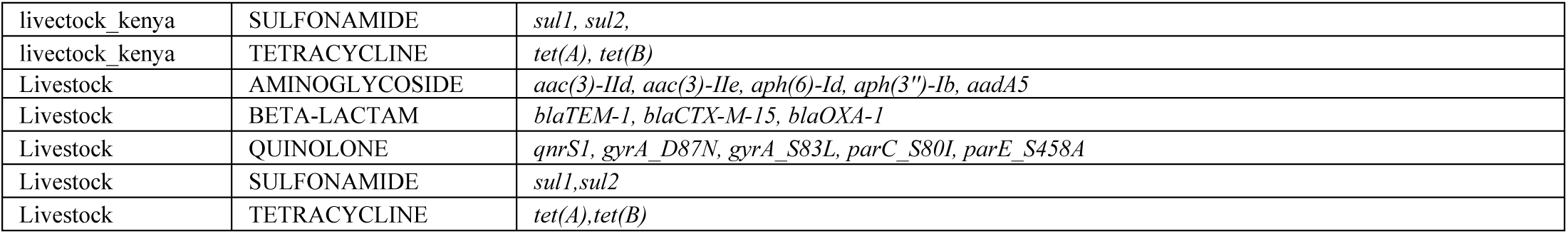
Table showing the distribution of detected resistance genes in selected antibiotics among *E. coli* isolates from humans, livestock, fish, and the environment.

### Heatmap and hierarchical Clustering of AMR Profiles

A Heatmap of the presence and absence of key AMR genes across the sample set (**Figure 1**), showed the hierarchical clustering of isolates based on their resistance profiles reveals distinct groupings that align with their sources. Isolates from clinical settings form a separate cluster from those obtained from environmental and animal sources, suggesting source-specific patterns in the acquisition and dissemination of resistance genes. Additionally, isolates from livestock in Kenya cluster separately from other isolates, indicating potential geographic or host-specific variations in AMR gene distribution. These findings underscore the influence of both sample origin and geographical location on the structure of AMR profiles and suggest possible transmission pathways across different environments.

**Figure 1:**
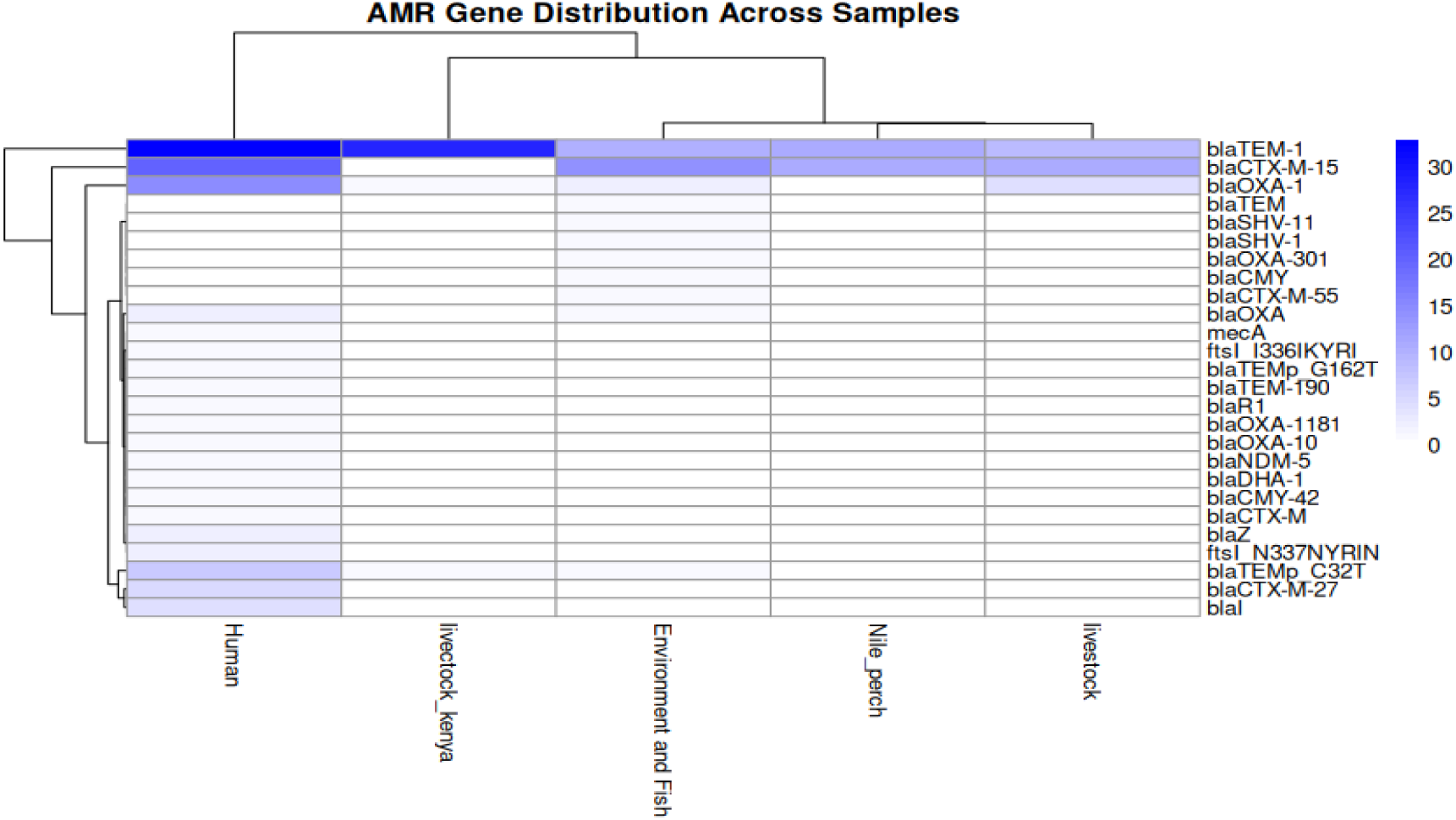
Heatmap plot of the relative abundance of ARGs in humans, livestock, Nile perch, environment and fish from Tanzania and livestock from Kenya. Clustered each group ARGs in rows columns and source in columns.

The Venn diagram illustrates the distribution of antimicrobial resistance genes (ARGs) across four ecological sources: humans, livestock (Tanzania and Kenya), fish, and the environment (**Figure 3**). The numbers within each circle represent the count of unique and shared ARGs across these sources, with percentages indicating their relative proportions. Human isolates show the highest number of unique ARGs (44, 40%), suggesting significant ARG diversity in human-associated *E. coli*. Environmental samples contain 16 unique ARGs (14%), reflecting the role of the environment as a reservoir for diverse resistance genes. Shared ARGs among the four sources are limited, with only 11 ARGs (10%) found across all groups, highlighting niche-specific genetic signatures.

**Figure 2:**
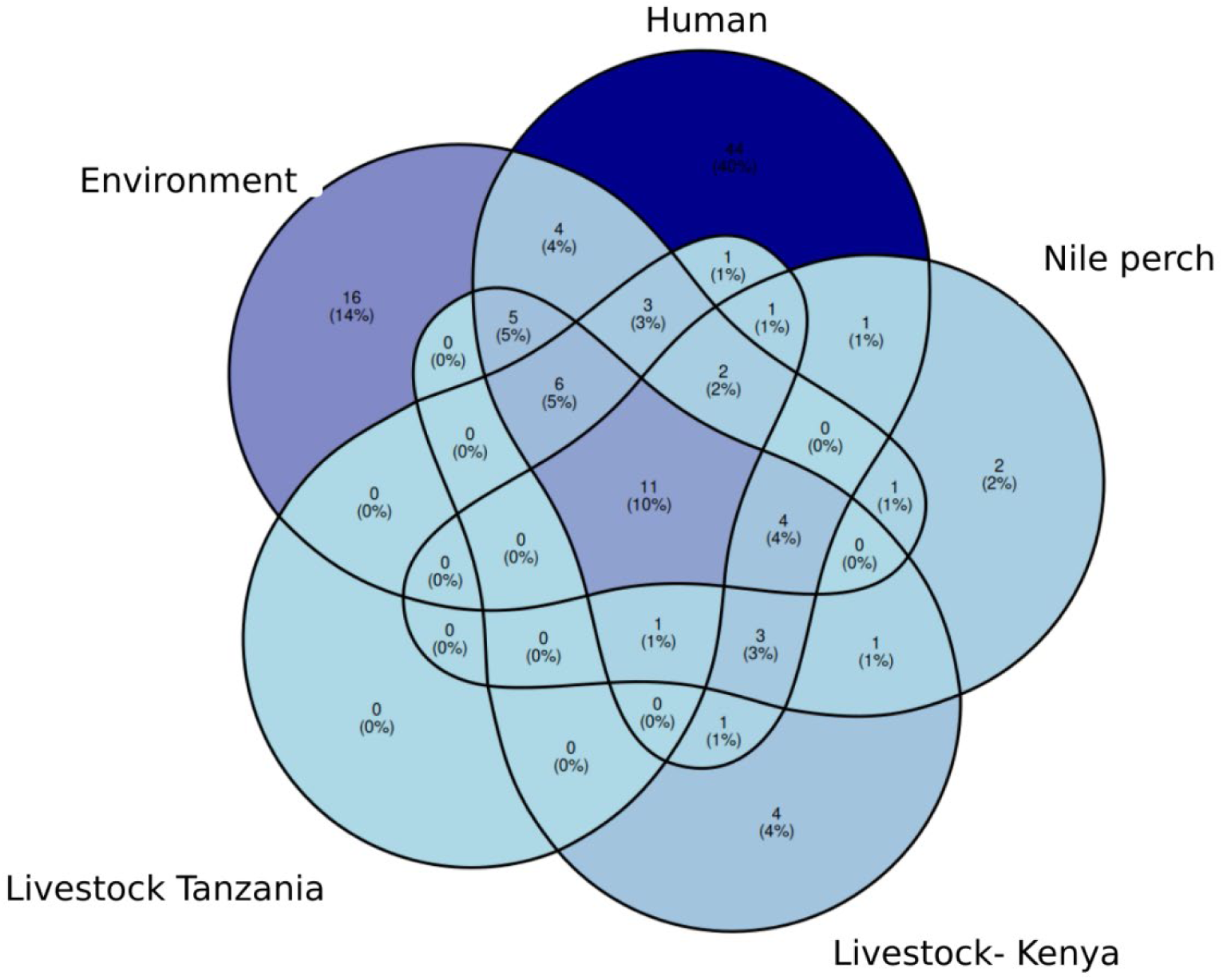
Venn Diagram of ARG distribution across different sources. Numbers in circles represent the count of unique and shared ARGs among sources, with percentages indicating relative proportions

**Figure 3:**
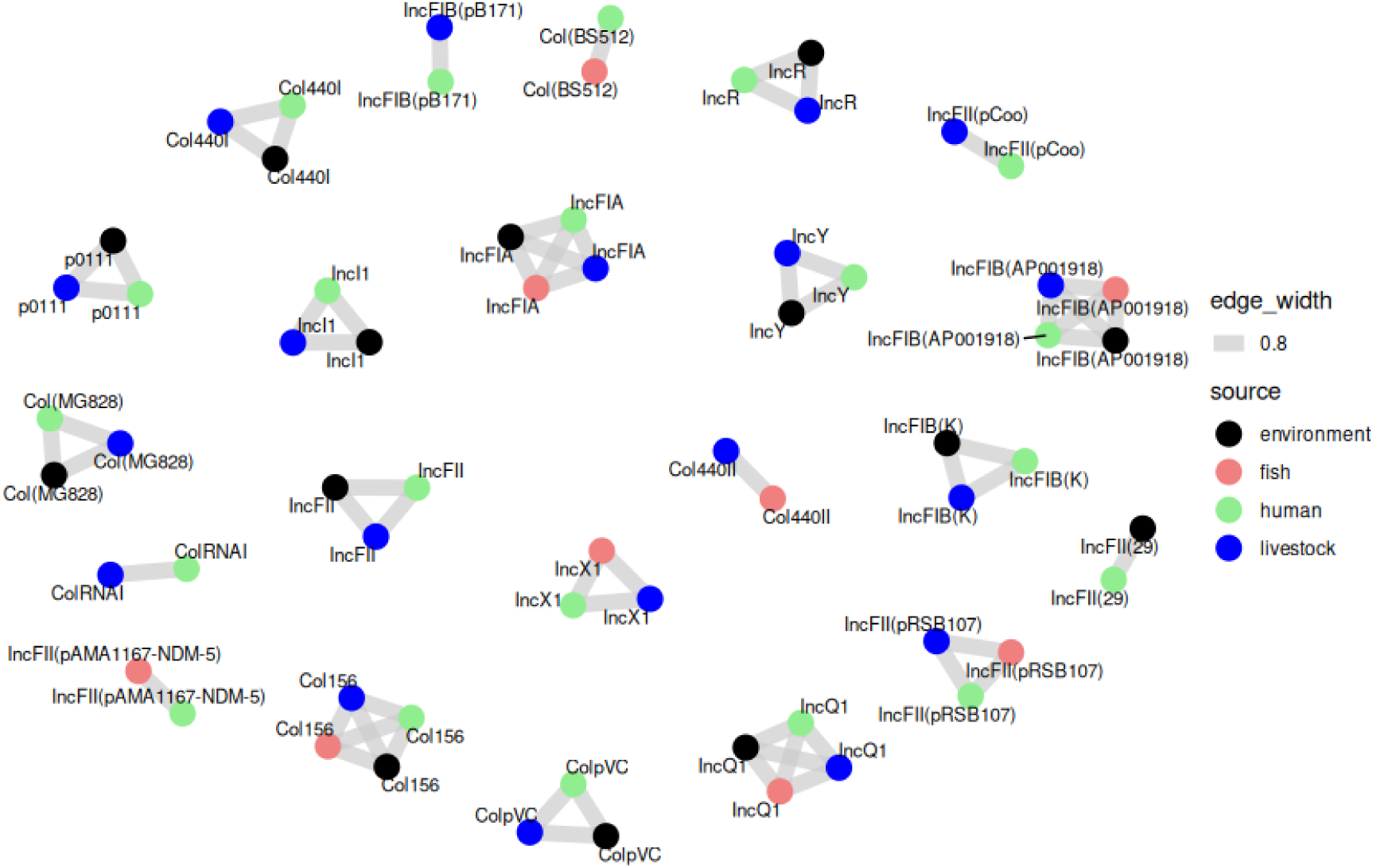
Plasmid Network of *E. coli* Isolates Across Humans, Livestock, Fish, and Environmental Sources. Which showed evidence of Cross-Source Horizontal Gene Transfer

#### The plasmid network

The plasmid network visualization depicts the relationships between plasmid replicon types detected in *E. coli* isolates from humans, livestock, fish (Nile perch), and the environment (**Figure 3**). Node colours represent sources (human = green, livestock = blue, fish = red, environment = black), while edges indicate shared plasmid types, with thickness reflecting connection strength. Plasmid types IncFIA, IncI1, and IncFII show the most extensive cross-source sharing, indicating their role in horizontal gene transfer. Human and livestock isolates exhibit the most significant connectivity, suggesting potential zoonotic transmission. Environmental nodes show widespread but weaker connections, reflecting their role as passive reservoirs.

### Population structure

The PCA analysis of *E. coli* isolates from humans, livestock, fish and the environment (**Figure 4**) showed isolates from Kenya (grey) form a tight and distinct cluster, possibly due to geographical factors or antibiotic use policies. Other isolates from humans, livestock, and the environment are more dispersed across the PCA space, implying greater genetic diversity. This variability may result from different ecological pressures, host-specific adaptations, and regional variations in antimicrobial use and environmental exposure. widely dispersed across the PCA plot, suggesting a higher degree of variability. This distribution highlights potential differences in *E. coli* populations across ecological niches and geographical regions. The observed patterns may provide insights into transmission dynamics, antimicrobial resistance dissemination, and environmental reservoirs of *E. coli* in Tanzania

**Figure 4.**
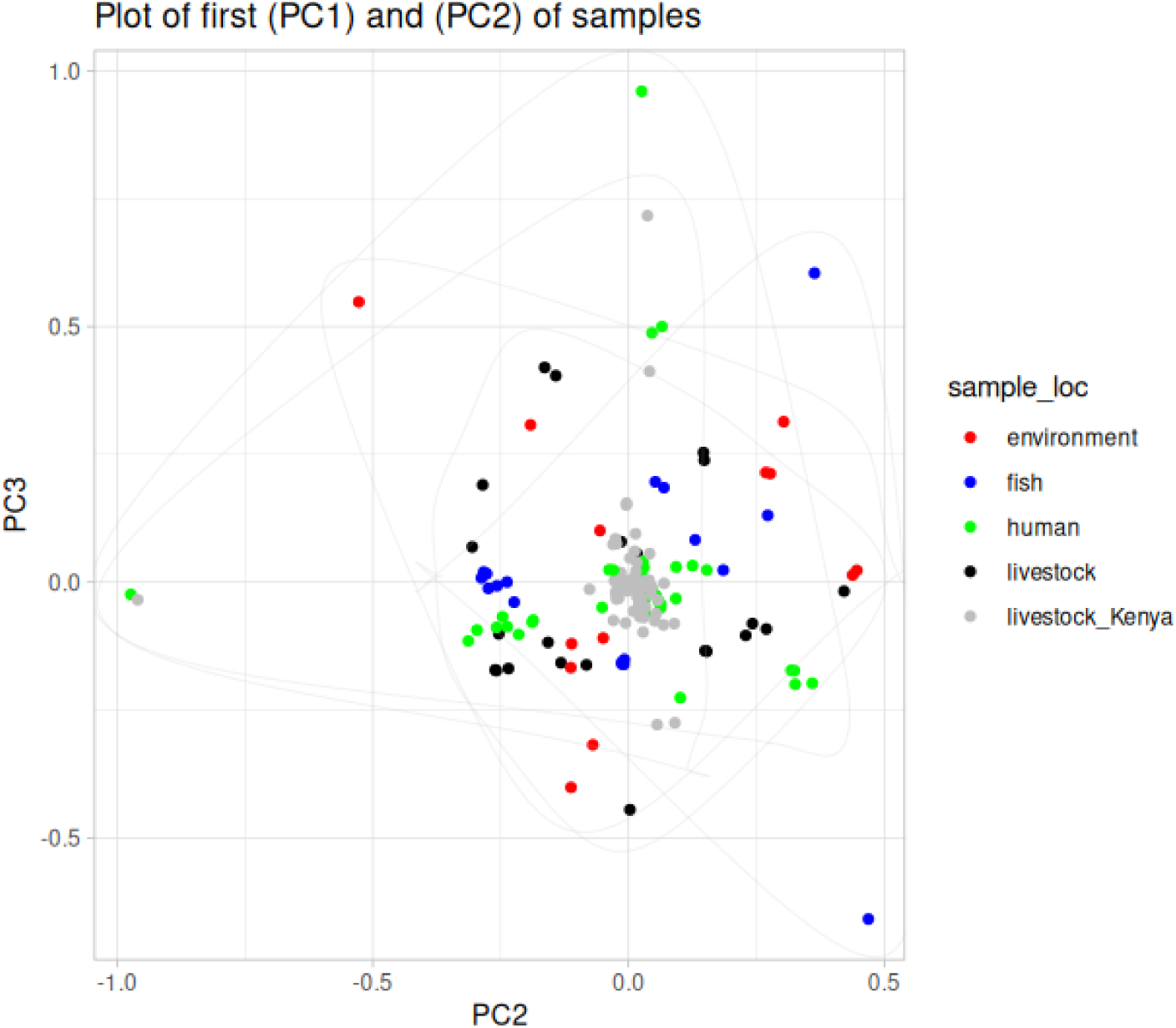
presents the principal component analysis (PCA) of *E. coli* isolates collected from various sources and locations. The isolates from Kenya (grey) formed a distinct cluster, indicating genetic or phenotypic similarities. In contrast, isolates from humans, livestock, and the environment were more.

### Minimum Spanning Tree of cgMLST

Results showed the pie chart which displays the proportion of ST from different sources. The sources include the environment, fish, humans, livestock, and livestock (Kenya). The distribution appears relatively even, with all sources contributing significantly. Livestock and human constitute a major proportion, highlighting their role in AMR transmission. Fish and environmental sources also contribute, reinforcing the multi-sectoral AMR dissemination. Fig 5 **(A).** Results showed also clustering patterns reveal that human isolates (red nodes) are closely related to livestock isolates (green, orange), suggesting potential transmission events. Environmental isolates (grey nodes) are distributed throughout the tree, implying their role as reservoirs or intermediates in pathogen transmission. Fish isolates (light blue) are interspersed, suggesting interactions between aquatic and terrestrial ecosystems. Fig 5 **(B)**

**Figure 5.**
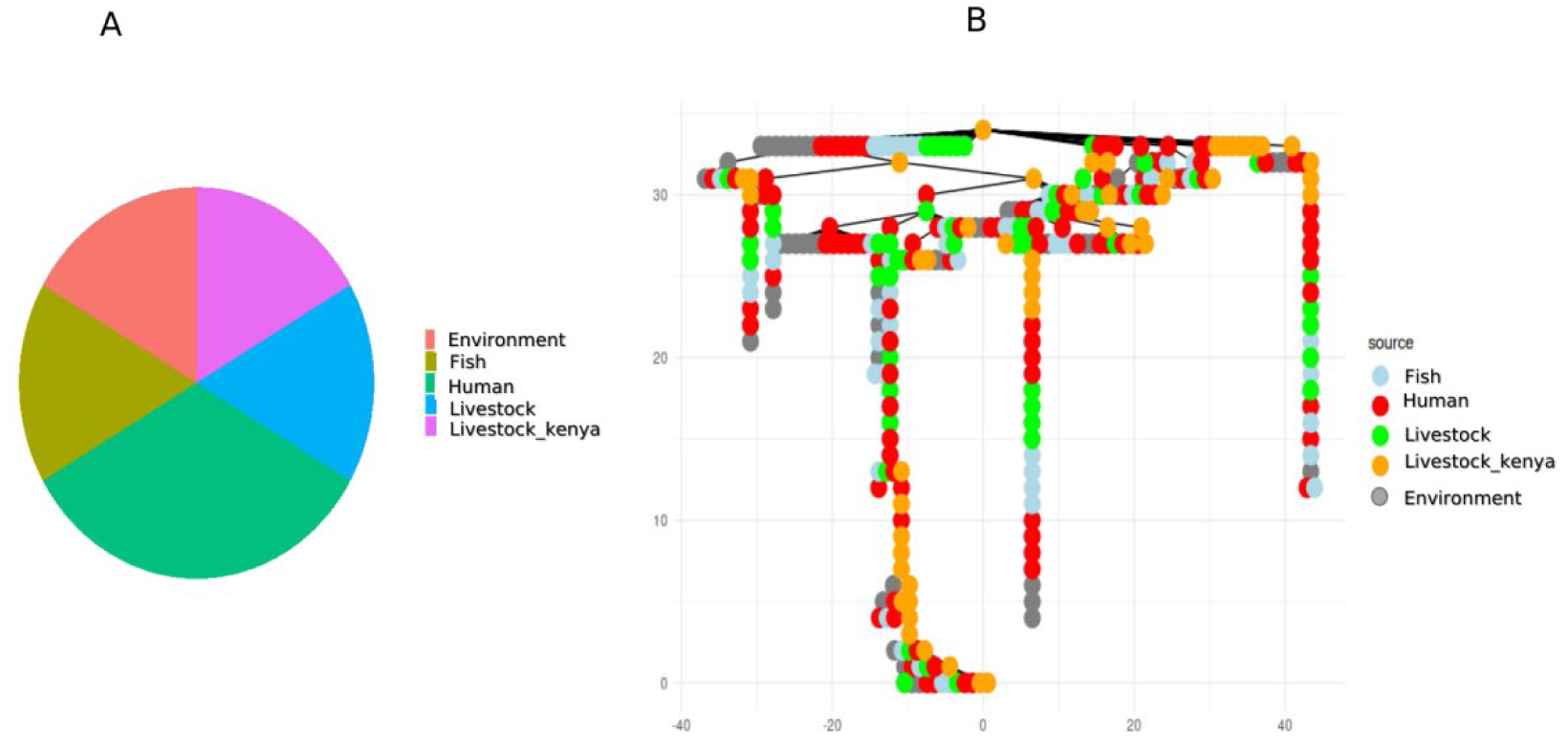
(A): The pie chart presents the proportional distribution based on allelic profiles from *E. coli* across different sources. (B) Minimum Spanning Tree (MST) based on core genome Multilocus Sequence Typing (cgMLST). Each node represents ST, and the edges indicate genetic relatedness. Nodes are colour-coded according to the source of isolation. The branching structure highlights potential evolutionary trajectories, with some clusters showing a dominant source while others appear mixed

## Discussion

Bacterial resistance toward broad-spectrum antibiotics has become a major concern in recent years. The One Health approach has been widely advocated to enhance the understanding of the transmission dynamics of antibiotic bacteria from different sources (Cella et al., 2023; McEwen & Collignon, 2018). Additionally, advancements in sequencing platforms and bioinformatics tools have significantly revolutionized the ability to trace AMR transmission between humans, livestock, and the environment (Quitmeyer, 2024; Satam et al., 2023). Furthermore, comparative analyses of different sequencing platforms and databases have been conducted to provide comprehensive analysis pipelines for defining AMR gene occurrence (Soni et al., 2021). These studies highlight the critical role of secondary data analysis in accurately identifying and characterizing AMR genes, thereby informing strategies to combat the spread of resistant pathogens. Collectively, these advancements demonstrate how secondary analysis of available sequence data could revolutionize our capacity to trace AMR transmission across various reservoirs. Therefore, this study leverages existing WGS data to elucidate the distribution and genetic relationships of antibiotic-resistant *E. coli* isolated from diverse sources within the One Health framework, encompassing humans, livestock, and environmental reservoirs, and the role of each source in the transmission of multidrug-resistant *E. coli*.

Results indicated a significant burden of antibiotic resistance genes across all sources. Among all antibiotic classes, aminoglycosides and β-lactams exhibited the highest prevalence across all samples. This finding is consistent with studies showing that resistance to β-lactams and aminoglycosides is increasingly common in human, animal, aquatic ecosystems and environmental reservoirs which, underlining the interconnectedness of human, animal, and environmental reservoirs in AMR transmission (Gaşpar et al., 2021; Gemeda et al., 2023; Kiiti et al., 2021; Pormohammad et al., 2019; Sonola et al., 2022). Compared to studies in sub-Saharan Africa, our findings align with reports from Tanzania and Kenya, which have documented high prevalence rates of ESBL-producing *E. coli* in livestock and humans (Kemp, 2020; Mwakyoma et al., 2023).

Notably, the blaCTX-M-15 gene was detected in nearly all samples, including isolates from fish and the environment. This gene encodes a β-lactamase enzyme that confers resistance to third-generation cephalosporins, particularly ceftriaxone, cefotaxime, and ceftazidime. Reports from other studies have reported the occurrence of bla_CTX*-M-15*_ in *E. coli* isolated from different sources ranging from humans, livestock and the environment (Lyimo, Buza, Subbiah, Smith, et al., 2016b; Minja et al., 2021; Moremi et al., 2016; Shawa et al., 2021). The widespread presence of bla*_CTX-M-15_* and aminoglycoside resistance genes suggests high levels of antibiotic pressure in both clinical and agricultural settings. The detection of these genes in environmental samples highlights the potential role of wastewater and agricultural runoff in spreading AMR. This is very alarming for the future of antibiotics under the cephalosporins group.

Resistance genes, such as *aph(3’’)-Ib*, *aph(6)-Id*, *aadA1*, and *aadA2*, which are responsible for resistance to the aminoglycoside class of antibiotics, particularly streptomycin, have been shown to exhibit high prevalence across all sources(Shi et al., 2013; Zhang et al., 2023). These resistance genes are important markers of the broader issue of AMR, which is increasingly complicating the treatment of infections caused by bacteria. The presence of these resistance genes in clinical isolates, as well as environmental and agricultural isolates, suggests a widespread dissemination of resistance mechanisms across various settings, including hospitals, communities, and agricultural environments.

In some isolates, the study found the occurrence of more than five different resistance genes occurring together. This contributes to the spreading of the genes to the environment including soil and water through manure back to humans by direct contact with farm animals, through exposure to animal manure, wastewater, or aerosol, and by consumption of uncooked animal products such as meat, eggs, milk, etc (Jaja et al., 2020; Zhang et al., 2023). This information is very critical in applying one-health in the control of AMR transmission.

The Venn diagram results showed humans exhibit the highest number of unique antimicrobial resistance genes (44, 40%), suggesting that medical antibiotic use plays a dominant role in resistance selection. These resistance determinants could be driven by excessive antibiotic prescriptions, self-medication, or hospital-acquired infections. The resistance-determinant genes can find their way back to animals or the environment.

The environment category contains a substantial number of unique ARGs (16, 14%), emphasizing the importance of wastewater contamination, aquaculture antibiotic use, and the environmental persistence of resistance genes. Since many resistance genes in the environment are derived from human and livestock sources, pollution control and water treatment strategies are critical for AMR mitigation. The overlapping ARGs between this category and others suggest the potential for ARG spillover into aquatic ecosystems, contributing to horizontal gene transfer (HGT) between bacterial communities.

Livestock samples from Tanzania and Kenya exhibit fewer unique ARGs and relatively low overlap with Nile perch. This suggests that direct transmission between these reservoirs is limited. However, ARG exchange may still occur through shared environmental exposure, such as contaminated water sources, runoff from farms, or the use of animal waste in aquaculture. The presence of some common ARGs (e.g., 11 shared ARGs, 10%) indicates potential indirect transmission pathways. Differences between livestock in Tanzania and Kenya indicate potential geographical, management, and antibiotic use policy variations. These findings reinforce the need for harmonized antimicrobial use policies in East Africa, focusing on standardized surveillance and interventions across borders. They were further confirmed by heatmap results which illustrated the distribution of antimicrobial AMR genes across various sample sources, including livestock from Kenya, human, livestock, fish and environmental (water and soil) and fish samples from Tanzania. The hierarchical clustering provides insights into the similarities between AMR gene profiles across sample categories and the co-occurrence of resistance genes. This result aligns with other studies which showed the co-occurrence of antibiotic-resistance genes across different sources (Altayb et al., 2022; Inda-Díaz et al., 2023). The highest AMR gene abundance was observed in human samples, particularly for aminoglycoside, beta-lactam, and quinolone resistance genes. The observed result aligns with previous findings where human-associated bacteria harbour a diverse and extensive repertoire of resistance genes, likely due to frequent antibiotic exposure in clinical and community settings.

The plasmid network visualization reveals important insights into the dynamics of horizontal gene transfer (HGT) in *E. coli* populations from different sources, including humans, livestock, fish, and the environment. The plasmids *IncFIA*, *IncI1*, and *IncFII* are notably widespread across multiple sources, indicating their pivotal role in facilitating horizontal gene transfer in *E. coli*. These plasmids are known to carry genes related to antibiotic resistance, virulence factors, and other traits that enhance bacterial survival (Chen et al., 2024; Lyimo, Buza, Subbiah, Temba, et al., 2016; Pankok et al., 2022; Rozwandowicz et al., 2018a, 2018b). Their extensive sharing between isolates from diverse sources suggests that they serve as key vectors for gene transfer, allowing for the spread of potentially harmful traits across species boundaries. The high connectivity between human and livestock isolates observed in the plasmid network suggests a significant role for zoonotic transmission in the spread of resistant *E. coli* strains. This sharing of plasmids has also been observed in other studies. For example, a study by Ibekwe *et al*. showed that *E. coli* isolated from swine and dairy manure carried plasmids such as IncFIA(B), IncFII, IncX1, IncX4, and IncQ(Ibekwe et al., 2021). The frequent interactions between human and livestock plasmid types imply that certain practices in livestock farming, such as the use of antibiotics, could be facilitating the spread of resistant strains. Additionally, the proximity of humans to livestock— whether through direct contact, consumption of contaminated meat, or environmental contamination might create opportunities for the exchange of plasmids carrying resistance genes (Graham et al., 2019; Velazquez-Meza et al., 2022). This is particularly concerning when considering the role of livestock as a reservoir for pathogenic bacteria that can affect human health. Furthermore, the presence of IncFIA, IncI1, and IncFII plasmids in both humans and livestock suggests that *E. coli* may be evolving to adapt to the selective pressures exerted by human and animal health systems. Understanding these zoonotic transmission pathways (Koutsoumanis et al., 2021) is essential for developing more effective public health policies and strategies for controlling the spread of resistant *E. coli*.

The environmental nodes in the network could represent passive reservoirs where *E. coli* strains persist in a dormant state or without direct pathogenic impact but contribute to the continuous introduction of plasmid-borne genes into different ecological niches (Koutsoumanis et al., 2021). Additionally, environmental conditions like contamination from agricultural runoff or sewage could lead to the spread of resistant strains in the broader ecosystem, indirectly affecting both animal and human populations (Koutsoumanis et al., 2021; Pormohammad et al., 2019). The use of plasmid network visualization provides a powerful tool for understanding the complex relationships between *E. coli* isolates from different sources.

The PCA results offer valuable insights into the complex interactions between humans, livestock, and the environment, particularly concerning *E. coli* transmission and AMR dynamics (McEwen & Collignon, 2018; Velazquez-Meza et al., 2022). The distinct clustering of the Kenyan isolates suggests that these isolates share a common genetic signature, possibly reflecting specific local factors that influence *E. coli* populations, such as regional environmental conditions, agricultural practices, or unique microbial reservoirs. The broader dispersion of the isolates from humans, livestock, and the environment across the PCA plot indicates significant variability within these groups, pointing to a heterogeneous nature of *E. coli* populations in different ecological niches. This could reflect a variety of factors, including the diversity of *E. coli* strains in different host species, environmental influences such as water quality or soil conditions, and the role of human behavior (e.g., antibiotic use, hygiene practices) in shaping microbial populations.

The results highlight the need for integrated One Health surveillance strategies to monitor and mitigate the spread of AMR across human, animal, and environmental reservoirs. The presence of clinically relevant resistance genes in non-human sources raises concerns about zoonotic transmission and the role of environmental compartments as AMR reservoirs.

### Conclusion and recommendations

This study highlights the extensive distribution of AMR genes across multiple ecological niches, emphasizing the urgent need for a coordinated One Health response. Key resistance genes were detected across all sources, with plasmid types IncFIA, IncI1, and IncFII showing extensive cross-source sharing, suggesting their critical role in facilitating horizontal gene transfer. Strong connectivity between human and livestock isolates indicates potential zoonotic transmission. PCA revealed a distinct cluster of Kenyan isolates, likely driven by localized antibiotic use and environmental conditions, while other isolates were more dispersed, reflecting greater genetic diversity influenced by different ecological and host-specific pressures.

Addressing these challenges requires a multidisciplinary approach integrating enhanced surveillance such as establishing regional AMR surveillance networks integrating human, veterinary, and environmental sectors. improved wastewater management, antimicrobial stewardship, and advanced genomic and bioinformatics techniques. Implementing these strategies within the One Health framework is essential to mitigate the spread of antimicrobial resistance across ecosystems. Future research should include targeted epidemiological studies to directly link transmission pathways and assess the impact of antimicrobial stewardship interventions.

#### Study Strengths and Limitations

This study leveraged publicly available datasets compiled from multiple independent studies, enabling access to a large and diverse sample set. While this approach enhances the breadth of analysis and potential for general insights, it also introduces variability due to differences in sampling frames, definitions, and methodologies used across the original studies. Care was taken to account for these differences during analysis; however, some residual heterogeneity may influence data comparability and interpretation.

## Data Availability

All raw WGS data, along with the reference E. coli strain K-12 substr. MG1655 were retrieved from the NCBI GenBank database using the following accession numbers: PRJEB12361, PRJEB12376, PRJEB71714, PRJEB32607

https://www.ncbi.nlm.nih.gov/bioproject/PRJEB12361/

https://www.ncbi.nlm.nih.gov/bioproject/?term=PRJEB12376

https://www.ncbi.nlm.nih.gov/bioproject/?term=PRJEB71714

https://www.ncbi.nlm.nih.gov/bioproject/PRJEB32607/

## Author Contributions

BL and VS conceived and designed the study. BL analyzed and interpreted the data. BL and VS wrote the manuscript, and submitted it for publication. The authors are driven by a commitment to advancing the integration of genomic surveillance into routine healthcare systems.

## Acknowledgements

The authors acknowledge all research teams and study participants involved in generating the Whole Genome Sequences and submitting them to public repositories.

## Ethical Clearance

No ethical clearance was required, as the data were downloaded from the public database.

---

**S1:**
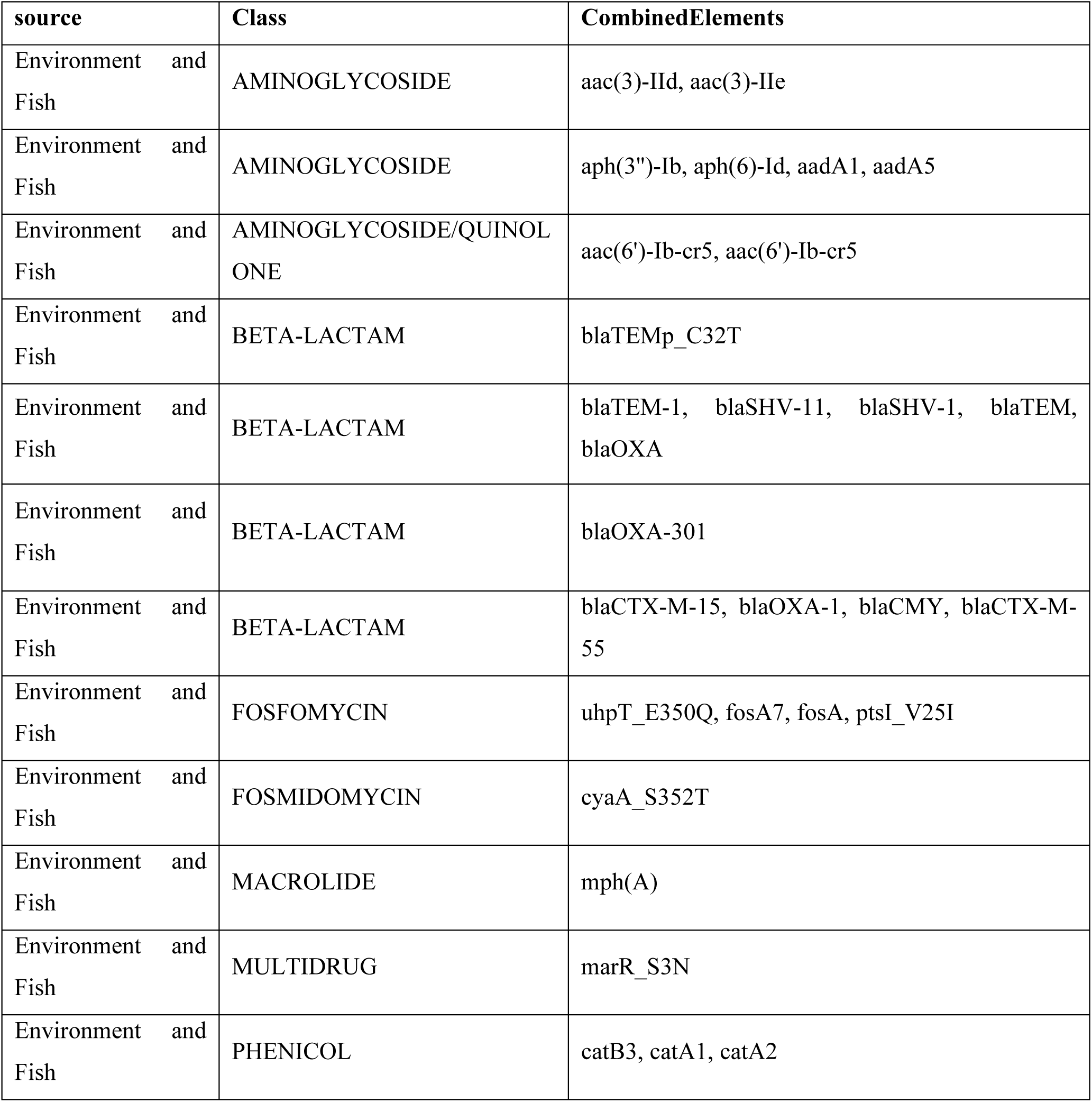
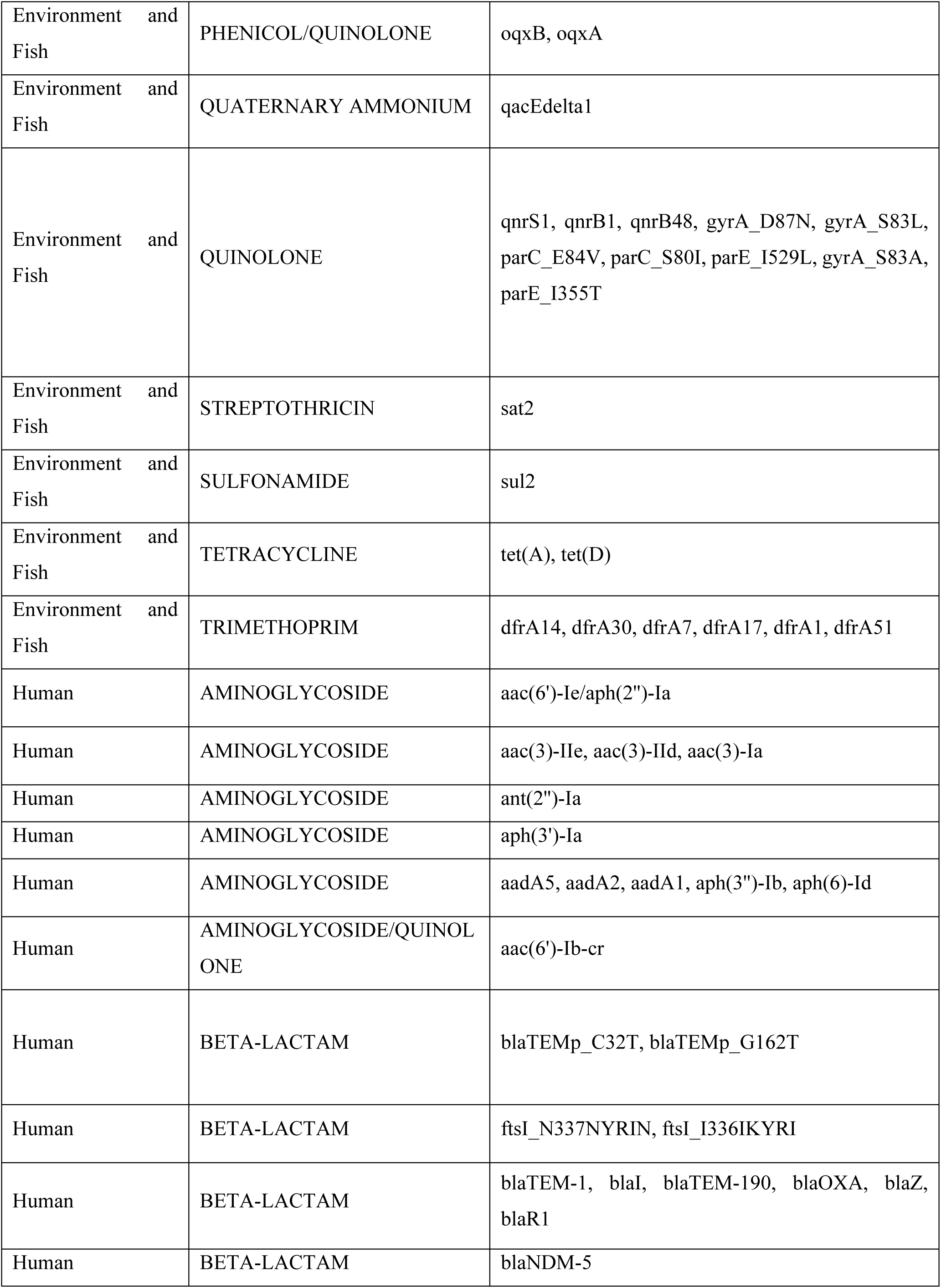
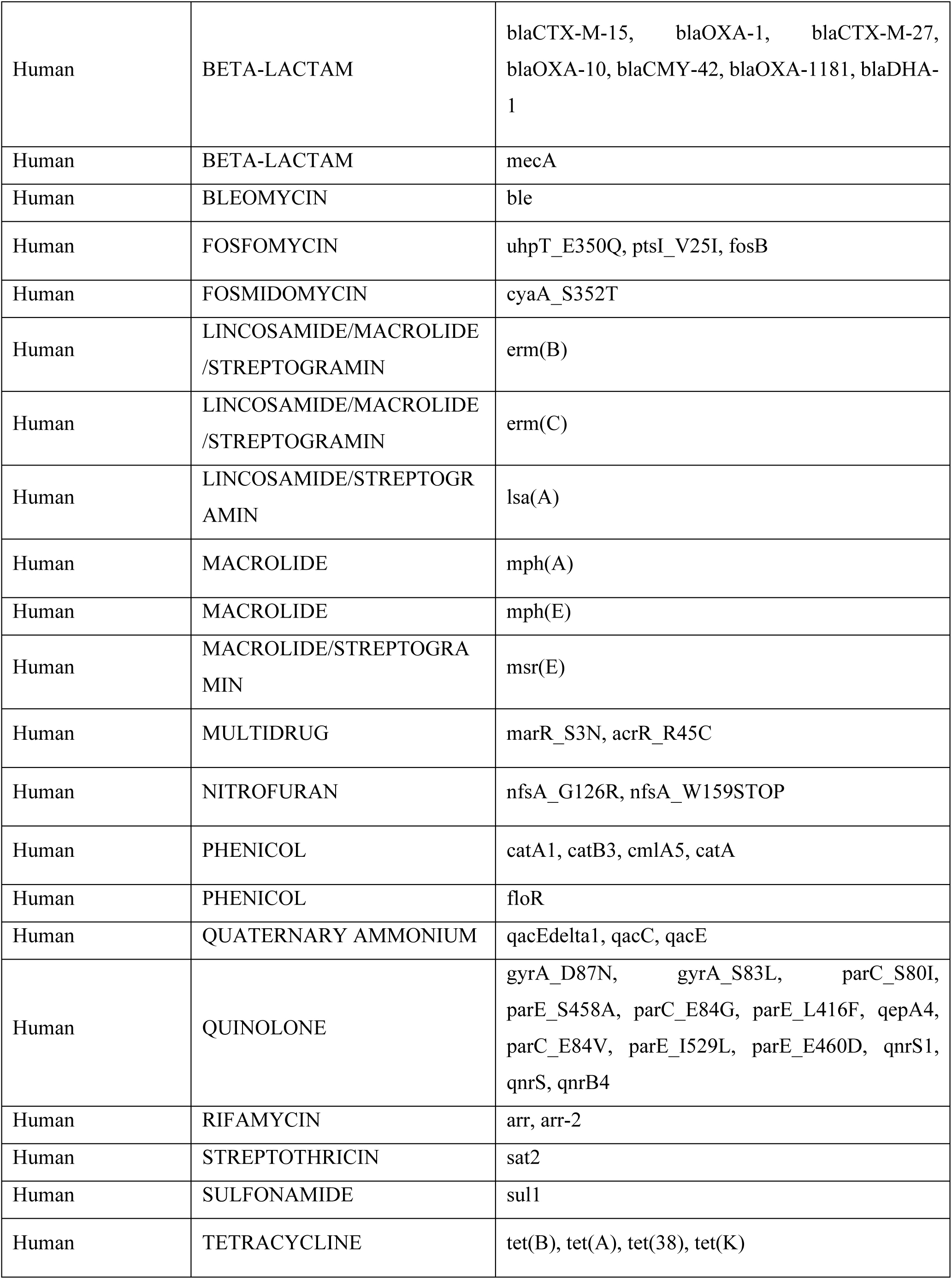
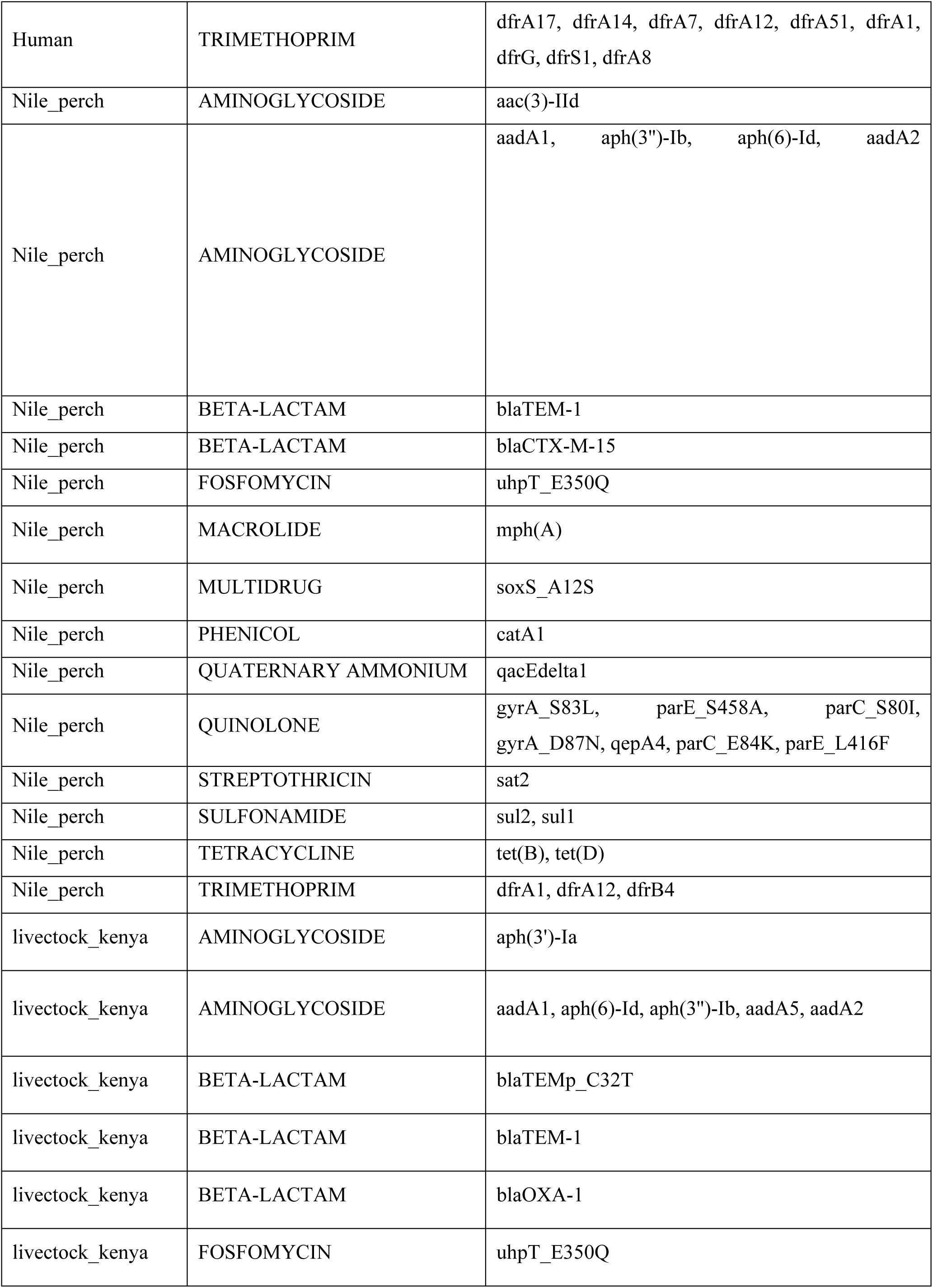
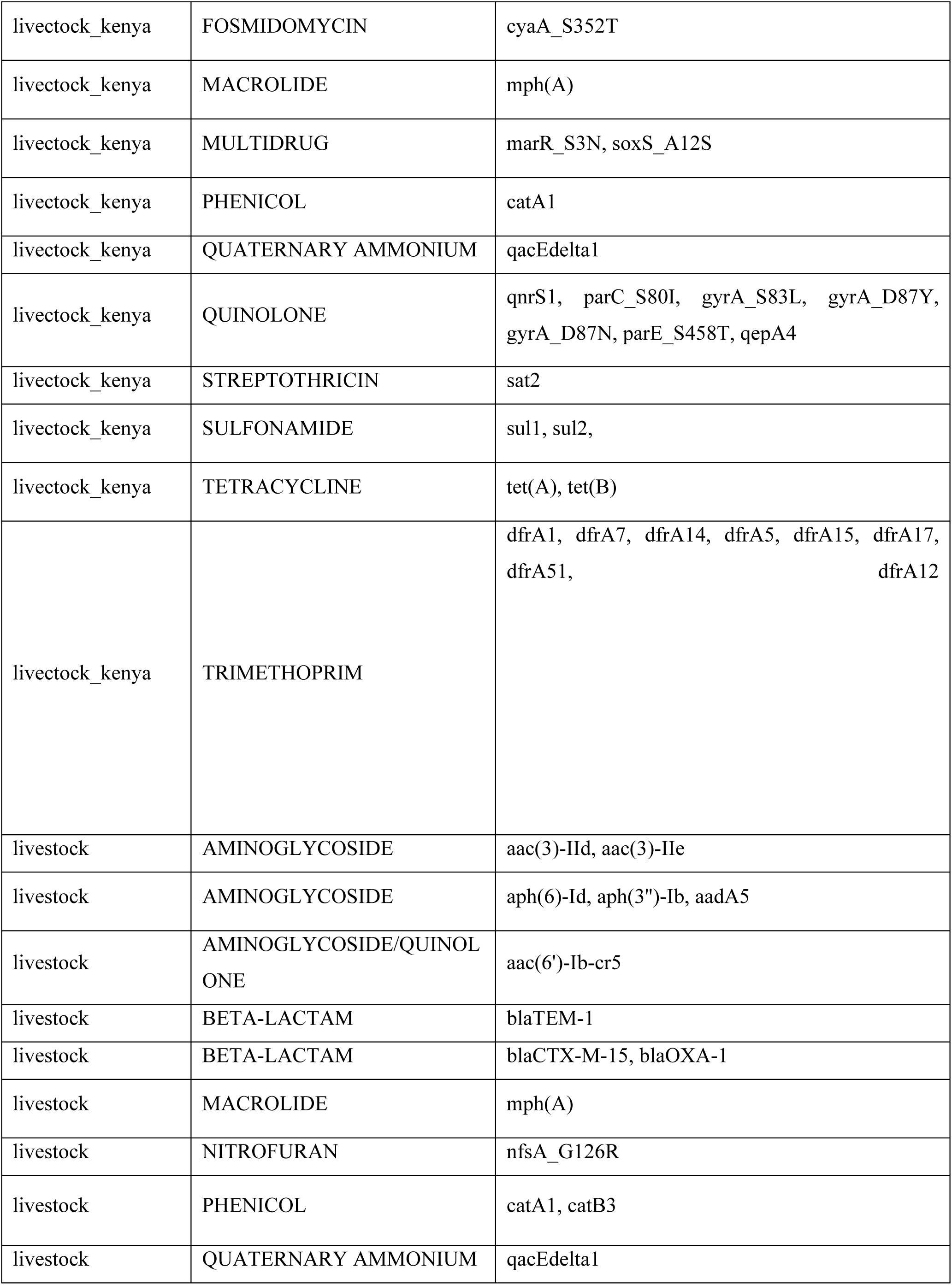
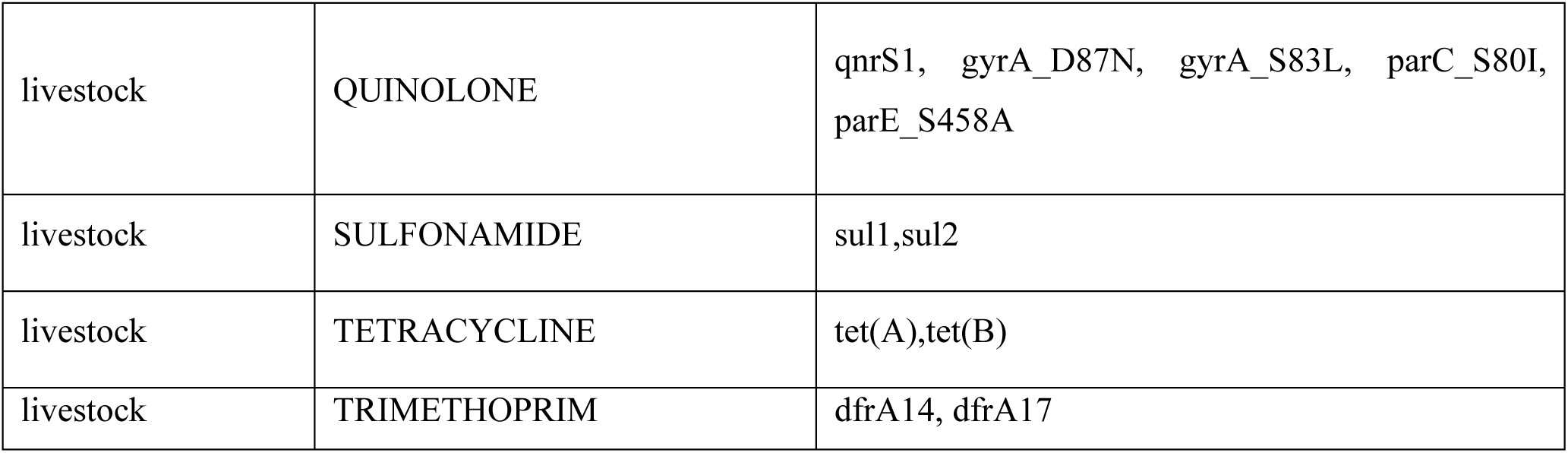
Supplement materials of Antimicrobial resistance (AMR) genes detected.

